# Transcriptomic Fingerprint of Bacterial Infection in Lower Extremity Ulcers

**DOI:** 10.1101/2021.12.20.21267962

**Authors:** Blaine Fritz, Julius Bier-Kirkegaard, Claus Henrik Nielsen, Klaus Kirketerp-Møller, Matthew Malone, Thomas Bjarnsholt

**Author notes:** **Corresponding Author:** Thomas Bjarnsholt.

## Abstract

Clinicians and researchers utilize subjective classification systems based on clinical parameters to stratify lower extremity ulcer infections for treatment and research. This study compared clinical infection classifications (mild to severe) of lower extremity ulcers (n = 44) with transcriptomic profiles and direct measurement of bacterial RNA signatures by RNA-sequencing. Samples demonstrating similar transcriptomes were clustered and characterized by transcriptomic fingerprint. Clinical infection severity did not explain the major sources of variability among the samples and samples with the same clinical classification demonstrated high inter-sample variability. High proportions of bacterial RNA, however, resulted in a strong effect on transcription and increased expression of genes associated with immune response and inflammation. K-means clustering identified two clusters of samples, one of which contained all of the samples with high levels of bacterial RNA. A support vector classifier identified a fingerprint of 20 genes, including immune-associated genes such as *CXCL8, GADD45B*, and *HILPDA*, which accurately identified samples with signs of infection via cross-validation. This suggests that stratification of infection states based on a transcriptomic fingerprint may be a useful tool for studying host-bacterial interactions in these ulcers, as well as an objective classification method to identify the severity of infection.

**Significance Statement:** Clinicians and researchers utilize classification schemes based on clinically measurable parameters to describe infection severity in lower extremity ulcers. However, here we show that the local host gene expression is often discordant to clinical classification scores. We observed this inconsistency is explained by the increased presence of bacteria, which promotes increased immune and inflammatory responses. Two groups of host gene expression, predominantly differentiated by the levels of bacterial RNA, could be classified with less than 20 genes. These results provide significant insights into host response to bacterial infection where bacteria are directly observed, rather than implied from clinical observation, and illustrated the limitations of clinical observations to stratify lower extremity ulcers.

## Introduction

Lower extremity ulcers present both humanistic and economic burdens to society. A UK study identified a prevalence of chronic lower extremity wounds in 6% of the population with management costs amounting to £328.8 million[1]. One type of lower extremity ulcer with high burden are diabetic-related foot ulcers (DFU). DFUs arise due to compromised arterial circulation, peripheral neuropathy, and repeated injury. Many individuals also demonstrate dysregulation of the immune response and poor glycemic control. The combination of these complex comorbitities likely increases the potential of DFUs to develop infection and osteomyelitis [2, 3]. More than 50% of DFUs will become infected [4], which increases the likelihood of poor clinical outcomes, risk of hospital admission, and lower extremity amputation [5, 6]. DFUs may take weeks, months, or years to heal and 65% of patients develop a new ulcer within 5 years [3, 7]. These cycles of re-ulceration and/or infection contribute to reduced quality of life, increased morbidity and mortality. Similarly, persons with leg ulcers can experience prolonged pain, social challenges, and decreased psychological well-being [6, 8-10].

Several classification systems exist for the stratification of DFUs and grading of infection severity. These include the Infectious Diseases Society of America/International Working Group on the Diabetic Foot (IDSA/IWGDF) guidelines [11], Wagner classification system [12], University of Texas system (UT) [13], site/ischaemia/neuropathy/bacterial infection/depth (SINBAD) system [14], diabetic ulcer severity score (DUSS) [15], and perfusion/extent/depth/infection/sensation (PEDIS) systems [16]. These classifications combine clinical data and observations to stratify wounds, classify infections, guide treatment, and predict outcomes [17]. A limitation of classification systems is that clinical observations are open to observer interpretation and may vary greatly, depending on the observer and/or the patient’s physiological state[18]. Underlying comorbidities such as diabetes mellitus or peripheral arterial disease further compound the clinical picture and have been implicated as causes of immune dysfunction and/or reduced infection symptoms [19, 20].

Recent advances in RNA sequencing technologies have allowed high-resolution examination of gene-expression of ulcer tissue and infecting bacteria at the bulk and single-cell level. For example, stratification by infection severity demonstrated increased microbial diversity as well as a unique host-response in severe DFIs [21, 22]. Wound healing has also been studied as an important outcome. Dysregulation of major transcriptional networks, such as those associated with migration of neutrophils and macrophages and inflammation have been identified in non-healing wounds [23, 24]. Unique subpopulations of fibroblasts have also been linked to improved wound healing [23]. Furthermore, transcriptomic studies have identified unique transcriptome of bacteria during chronic infection [25]. Studies examining transcription as a factor of infection severity often utilize clinical scores to classify highly infected samples, though there is evidence that clinical infection scores do not correspond with bacterial load [18].

In this study, we analyzed RNA sequencing data of clinically infected DFU biopsies to examine the effect of bacterial load on host transcription and correlation to clinical parameters. We assessed both the clinical infection severity score (IDSA) as well as direct measurement of bacterial activity in the samples to explain changes in the host transcriptome. We then identify differentially expressed genes and pathways which are characteristic for DFUs demonstrating signs of bacterial infection. From these genes, we develop a transcriptomic fingerprint of ∼20 genes, which could be utilized to identify infected DFUs and guide treatment.

## Methods

### Sample collection and external data sources

Tissue biopsies were collected from either the Liverpool Hospital (LHS) High-Risk Foot Service, Liverpool Hospital, Liverpool, Australia. External RNA-sequencing data utilized for the fingerprint validation was obtained from Heravi et al[22]. LHS samples were collected from patients presenting with an infected foot ulcer, where a punch-biopsy was taken post-debridement. Detailed tissue collection methods are described in Supplementary Information (SI). All tissue samples were immediately placed into RNAlater, incubated at 4°C for 24 hours, and then frozen at -80°C until RNA extraction. The clinical metadata categories of interest were ulcer duration (0 = less than 2 weeks, 1 = 2 to 4 weeks, 2 = greater than 4 weeks), PEDIS/IDSA infection score (2 = mild, 3 = moderate, 4 = severe) [38].

### RNA extraction, library preparation, and sequencing

Frozen samples in RNAlater were thawed on ice. The tissue was removed and placed inside an empty, sterile Petri plate. A scalpel was used to cut a piece of tissue with a volume of approximately 0.5 cm^3^. RNA extraction was then performed chloroform/phenol phase separation, as previously described [25, 39] with some modifications. The tissue was placed in a 2 mL microtube (Sarstedt, Germany, Cat: 72.693.005) filled one-third with 2mm and 0.1 mm diameter zirconia beads (Biospec, USA). One milliliter of RNABee (Amsbio, UK, cat: CS-501) containing 10 µL/mL β-mercaptoethanol was added to the tubes. The tubes were placed in a MagNA Lyzer instrument (Roche) and homogenized at maximum power for 3 × 30 seconds. Tubes were placed on ice for 1 min after each homogenization step. The tubes were shaken vigorously for 30 sec, incubated for 5 min on ice, and then centrifuged at 13,000 x g for 30 min at 4 °C. The upper aqueous phase was collected and placed in a new, 1.5 µL DNA Lo-Bind^®^ centrifuge tubes (Eppendorf AG, Germany, cat: 0030108051). Ice-cold ethanol (0.5mL) and 2 µl of 5 mg/mL linear acrylamide was added. The tubes were inverted several times and stored at - 80 °C overnight. The samples were then thawed on ice and centrifuged (13,000 x g, 30 min, +4 °C). The supernatant was discarded and the pellet was washed twice with fresh, ice-cold 75 % ethanol. The pellet was then resuspended in 20 – 65 µL nuclease free water. RNA concentration was quantified with a NanoDrop spectrophotometer. Contaminating DNA was removed by combining ∼2.5 µg RNA with 3 µL RQ-1 RNAse Free DNase (Promega, USA), 3 µL DNAse buffer solution, 1 µL RiboGuard™ RNAse inhibitor (Lucigen, USA, cat: RG90925), and nuclease free water to a final volume of 30 µL. The DNAse-treated RNA was then re-purified with the RNABee protocol described above and then stored at –80°C. Ribosomal RNA depletion was performed either with the 10:1 human:pan-prokaryote riboPOOLs (siTOOLS Biotech, Germany, protocol version: 1.4.2). The NEBNext^®^ Ultra II RNA library preparation kit (New England Biolabs (NEB), USA, cat: E7775S) was used to prepare cDNA libraries for the LHS samples. Concentrations and quality of the final libraries were assessed with a Qubit 4 fluorometer and Agilent Bioanalyzer. Samples were sequenced either on an Illumina HiSeq4000 or Novaseq sequencing instrument.

### QC processing, alignment, quantification, and bias control of sequence data

All raw sequence data, including external data, was processed at the same time, using the same pipeline, though accounting for paired-end or single-end data. Adapter and quality trimming was performed with cutadapt 2.4 [40]. Reads less than 20 nucleotides after trimming were discarded. In-silico rRNA depletion was performed with SortMeRNA v 2.1 against the SILVA rRNA database to remove eukaryotic, bacterial, or archaeal ribosomal sequences. The reads were then aligned with bwa-mem with default settings (v. 0.7.16a) against the GRCh38 human genome assembly (GCA_000001405.15, RefSeq, full analysis set), including all alternative haplotypes and unlocalized scaffolds. Aligned reads were then assigned to exon features in the using the NCBI RefSeq annotation with featureCounts [41]. Any samples containing less than 1 million reads then discarded from the analysis. Any exonic features which represented other transcript types, such as ncRNA and tRNA, were also removed from the analysis. To normalize for batch differences between data sets, differential gene expression was performed between LHS data and the external data from Heravi et al [22]. Any genes identified as differentially expressed in this analysis were removed from the data set. This data was then used for further analysis.

### Principal component analysis, k-means clustering, and differential gene-expression

The raw count data was normalized using DESeq2’s variance stabilizing transformation (vst, blind= TRUE, nsub = 1000) and analyzed with the prcomp function in R with no additional scaling. The first two principal components were plotted (Figure 1). Component loading analysis and principal component correlations were performed with the R package, PCAtools. Spearman correlation with benjamini Hochberg correction for multiple comparisons was used for the correlation analysis of metadata with principal component positioning. The optimal number of clusters for k-means was selected by the fviz_nbclust function the factoextra package (v. 1.0.7) using the “silhouette” method. Differential gene expression analysis was performed with DEseq2 (v. 1.28.1), using default settings. Samples missing data in any of these metadata categories included in the models were removed from the dataset before the analysis. To identify differentially expressed genes due to clinical parameters, the IDSA/PEDIS score, categorical ulcer duration and a binary variable for if the sample contained >10 percent bacterial reads was used for DEeq2 model formula. To identify differentially expressed genes due to k-means cluster, the only variable in the formula was k-means cluster. In all cases, genes with an adjusted p-value of < 0.05 and an absolute log2 fold-change >2 were considered significantly differentially expressed.

**Figure 1.**
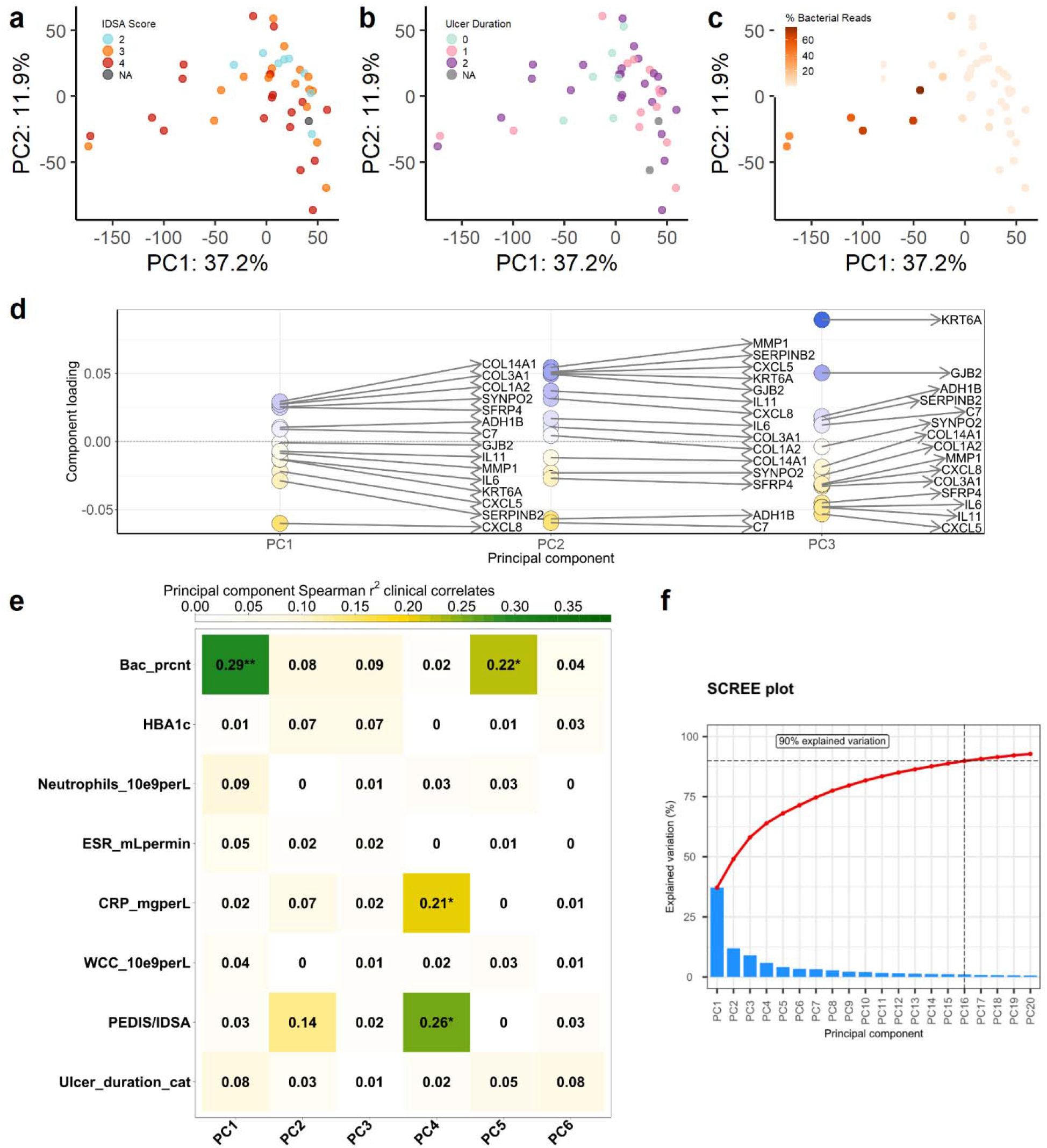
**(a-c)** Characterization of host gene expression ulcer transcriptomes (n = 12,378 genes) by principal component analysis. Points are colored by: **(a)** IDSA/PEDIS infection severity score [2:mild, 3:moderate, 4:severe], **(b)** Ulcer duration [0: less than 2 weeks, 1: 2 to 6 weeks, 2: Greater than 6 weeks], and **(c)** Percentage of all RNA-seq reads classified to bacteria. **(d)** Component loadings for the top 5% of positive and negatively weighted genes for PC1 to PC3. Points are shaded by component loading value. **(e)** Spearman correlation coefficients of metadata variables with positioning of a samples along PC1 to PC6. Significance tests were performed with benjamini-hochberg p-value correction for multiple comparisons [**: p<0.01, *: p<0.05]. (f) Scree plot demonstrating the percent of explained variance for PC1 to PC20. The red line represents the cumulative percentage of explained variance across these PCs

Functional characterization of the groups identified by k-means clustering was performed using PANTHER[42], using fisher’s exact test and false-discovery rate FDR < 0.05. Significantly differentially expressed genes with a positive log2 fold-change for a given comparison were used as the input and the list of all genes used as input to the differential gene expression analysis was used as the reference list. The annotations list used was the “GO biological process complete” data set (GO Ontology database DOI: 10.5281/zenodo.4081749, Released 2020-10-09).

### Feature Selection and Testing

To identify gene features that could be used to identify specific levels of clinical metadata or clusters of samples, a support vector machine (SVM) with a linear kernel with was fitted to the normalized data for PEDIS/IDSA score and k-means cluster. The most important features in the model were then selected based on the SVM coefficients to select for features for classification. To evaluate the optimal number of features to include in the model, this process was repeated for up to 100 features. For each number of features (1-100), the given number of features was selected and a stratified 6-fold cross validation was performed. Twenty genes were selected by the authors as the optimal number to avoid overfitting and account for additional variability when using external data. All analyses were performed with the python library scikit-learn (v. 1.0) [43].

## Results

RNA-sequencing was performed on thirty DFUr biopsies obtained from Liverpool Hospital, Liverpool Australia (samples annotated as P500-P529). Raw RNA sequence data (n = 14) from [22] was also included in the data set (samples prefixed with HH*). The combined data set yielded an average of 153 ± 23.9 M reads per sample passing quality filters. Samples from the Liverpool data showed significantly more rRNA contamination than the HH data with mean percentages of rRNA contamination of 57.4 ± 17 and 5.0 ±1, respectively. The non-rRNA reads were then aligned to the human reference genome and all reads mapping to exonic gene features were counted. Samples with less than 1M reads were excluded. Additionally, sample HH5 was excluded, as it was an extreme outlier. To control for variability due to source, differential gene expression analysis was performed, and 14,210 genes identified as differentially expressed were removed from the analysis, leaving 32,841 genes, which were analyzed (unless otherwise noted). The final data set contained a mean and median of 26.7 ± 21M and 20.9M reads per sample, respectively.

### Presence of bacteria is associated with shift in host transcriptome

To evaluate the effect of bacteria on the host transcriptome, the data was first normalized and summarized with principal component analysis (PCA). The results of the PCA analysis are plotted in Figure 1. The first two principal components summarized 49.1% of the overall variability. We observed that the majority of samples clustered positively along the first principal component (PC1), while a subset of samples was spread across PC1 in the negative direction.

The samples were evenly distributed along PC2. To determine which factors affected this positioning, we tested whether clinical metadata variables or percentage of bacterial reads in the samples correlated with any of the first 10 principal components. The proportion of bacteria/human reads was the factor showing significant correlation along either PC1 and PC2 (r^2^ = 0.29, p<0.05, spearman correlation with benjamini-hochberg correction). Levels of C-reactive protein as well as infection classification score showed slightly significant correlation to PC4, but this represented only a small proportion of the overall variability in the data. Additionally, the effect of increased proportions of bacterial reads was more prominent than batch variability between sources prior to correction of batch effects (Figure 1C, Supplementary Figure 1).

To further demonstrate that the positioning represented biological effects of bacteria rather than confounding factors, such as decreased sequencing depth of the host due to the high presence of bacteria, we examined which genes drive the variation across each principal component. Of the top 5% most weighted genes, the genes with the strongest loading in the negative directions included inflammatory cytokines (*CXCL8, CXCL5, IL6, IL1*), keratinocyte factors associated with bacterial infection (*KRT6A*) and a matrix metalloprotease induced under inflammation (*MMP1*). The genes with the strongest positive loading across included collagens associated with extracellular matrix deposition (*COL14A1, COL3A1, COL1A2*), actin-binding (*SYNPO2)*, apoptosis (*SFRP4)*, alcohol metabolization (*ADH1B)*, complement (*C7)*, and cellular gap junctions (*GJB2)*. This suggested that shift in positioning across PC1 in the negative direction is due to increased expression of immune-related genes, likely in response to bacterial infection.

### Enrichment of Immune Processes and Inflammation in Samples with High Bacterial Activity

To confirm that samples with increased bacterial activity represented a specific transcriptomic response and to generate groups for comparative analysis, we performed k-means clustering on the normalized expression data. This analysis identified two cluster of samples, C1 (n=8) and C2 (n=36) (Figure 2a). The mean proportion of bacterial:human reads per sample was significantly higher in C2 (t = 4.0367, p-value = 0.005, Welch t-test; Figure 2C). We then tested for genes that showed differential expression between these two clusters using DESeq2. This analysis identified 2793 genes differentially expressed (|log2FoldChange| > 2, adjusted p-value <0.05) between these two groups. Of these, 251 and 2541 genes showed significantly increased expression in C1 and C2, respectively. In comparison, the same analysis identified only identified 114 genes differentially expressed between different levels of ulcer severity, likely due to high variability among samples with the same clinical classification.

**Figure 2.**
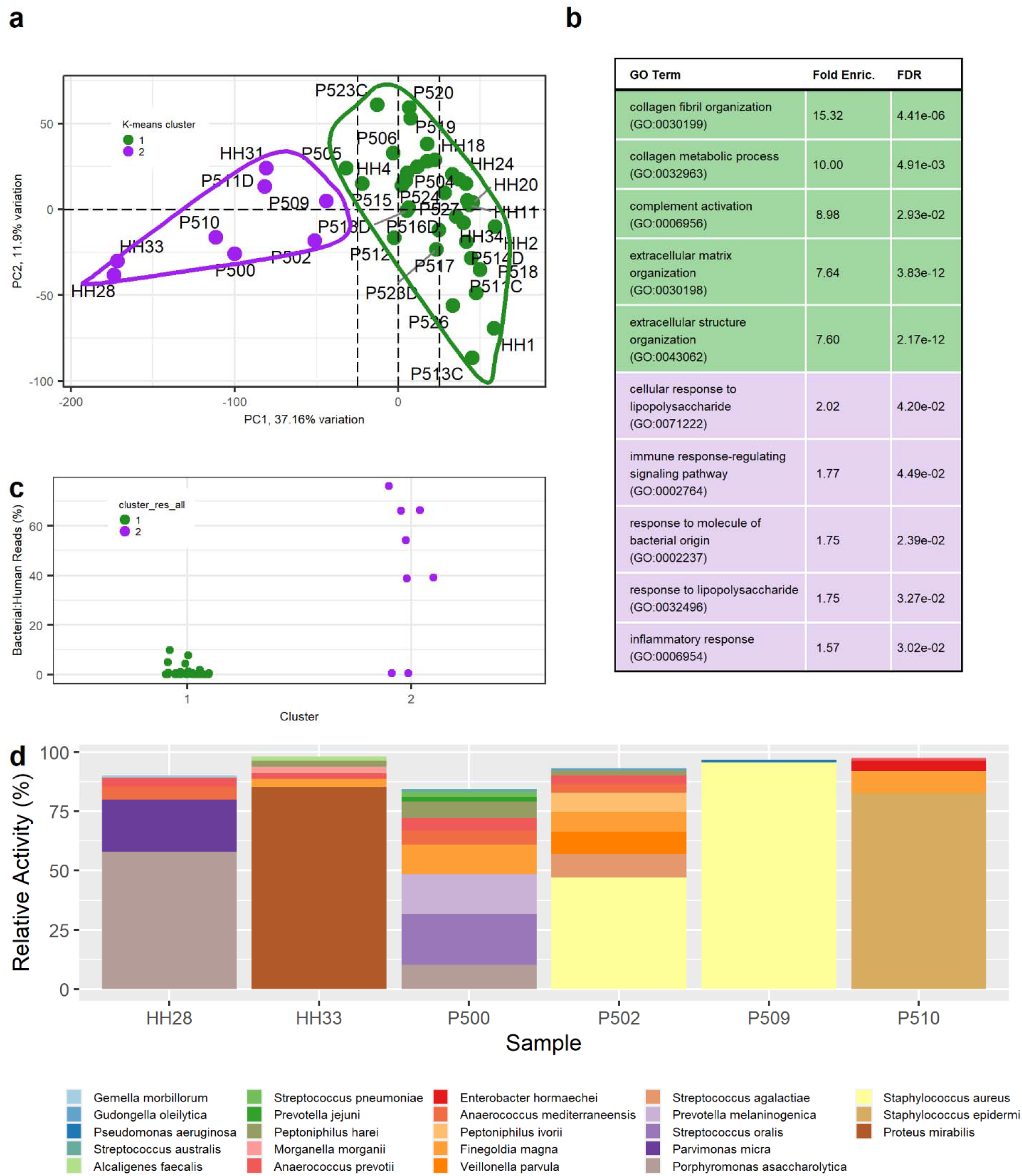
**(a)** Two clusters (C1 and C2) were identified by k-means analysis. Results are displayed projected over the principal component analysis plot of all genes (n = 12,378) with points colored by k-means cluster. Samples are labeled by ID, where samples prefixed with “P” and “HH” are from this study and Heravi et al [22], respectively. **(b)** Gene ontology (GO) terms for GO biological processes showing a significant overrepresentation (Fisher’s exact test) of genes identified as differentially expressed between C1 and C2. The top 5 overrepresented pathways for cluster 1 (green) and cluster 2 (violet) are shown. **(c)** Percentage of RNA-seq reads classified to bacteria relative to the total number of reads classified as either bacterial or host for C1 (green) and C2 (purple). The mean percentage of bacterial:human reads was significantly higher in C2 (t =4.04, p = 0.004, Welch t-test) **(d)** Relative activity (percentage of RNA reads for a specific species relative to all bacterial reads, %) for bacterial species with relative activity greater than 1%. Only samples with greater than 10% bacterial reads are shown.

C1 demonstrated increased expression of the cytokines (*CXCL12, CXCL13*) and cadherins *(CD34, CD36*). C2 demonstrated increased expression of *S100A8/9* and *S100A12. ADAM8* showed significantly increased expression in C2. In C2, we identified significantly increased expression of several leukocyte-associated cytokines (*CXCL8, CXCL2, CXCL16, IL6*, etc) and cadherins (*CD53, CD69*). We also observed increased expression of NFKB2, but also relatively higher expression NFKB inhibitors (*NFKBIA, NFKBIZ*). Though differentially expressed in C2, expression of TNF-alpha was low.

To examine whether the genes differentially expressed between the C1 (low bacterial activity) and C2 (high bacterial activity) represented enrichment of known biological processes, we performed a Gene Ontology enrichment analysis using PANTHER (Figure 2b). In the cluster with high bacterial activity (C2), we identified enrichment of 28 pathways (FDR<0.05), including the specific subclasses, “cellular response to lipopolysaccharide” (GO: 0071222), “immune response-regulating signaling pathway” (GO:0002764), “inflammatory response” (GO: 0006954), and “innate immune response” (GO: 0045087). In C1, there were 15 significantly enriched pathways (FDR<0.05), including “collagen fibril organization” (GO:0030199), “collagen metabolic process” (GO:0032963), “complement activation” (GO:0006956), “cell-matrix adhesion” (GO:0007160).

### Differential host response to Staphylococcus aureus

To investigate which bacterial species were present and active in the samples, KRAKEN was used to identify reads originating from bacteria and assign taxonomy (Figure 2d). In four of the six samples with high bacterial signals, at least 50% of the reads classified to bacteria were identified to a single species (*S. aureus, S. epidermidis, Proteus mirabilis*, or *Porphyromonas asaccharolytica*). We hypothesized that samples with increased signals of infection (i.e. C2) would display decreased alpha diversity. There was, however, no significant different in the number of species between the clusters (t = -0.071, p = 0.94, Welch t-test) for species with a minimum of 5% bacterial reads. We further investigated whether *Staphylococcus aureus*, which was highly active in two samples with increased signals of infection (C2) was also present in other samples with lower signals of infection (i.e. C1). Interestingly, *S. aureus*, was also found in 13 samples with low signs of infection (C1). This demonstrates a difference in host response between wounds, despite the presence of the same pathogenic bacterial species.

### Definition and validation of transcriptomic fingerprint to classify ulcer status

To identify a small set of genes, which could be used to identify samples with a bacterial infection, a support vector classifier (SVC) was applied to the RNA-seq data to select a reduced set of gene-features to define each cluster. Results of this analysis are displayed in Figure 3. Twenty gene-features were selected, which were selected from the model as useful classifiers to differentiate between samples in C1 and C2 (Figure 3a,b). We evaluated the accuracy of the classifier when trained with between 1 and 100 features and obtained high accuracy with less than 10 genes, but conservatively included 20 genes to increase the robustness of the model (Figure 3d). Several of the identified genes were associated with immune cells and inflammation, including *CXCL8* (neutrophil recruitment [26]), *GADD45B* (stress-response[27]), *HILPDA* (macrophage infiltration [28]), and *KIF21B* (T-cell polarization [29]). The normalized expression of these genes was also clearly elevated in C2 relative to C1 (Figure 3e). Genes that were negative classifiers for C2 (i.e. demonstrated increased expression in C1) included metalloproteases *MMP10* and *MMP12*, the collagen matrix protein *COL1A2*, and the chemokine *CCL21*. The feature with the largest coefficient was *SLCO2A1*, which showed increased expression in C1.

**Figure 3.**
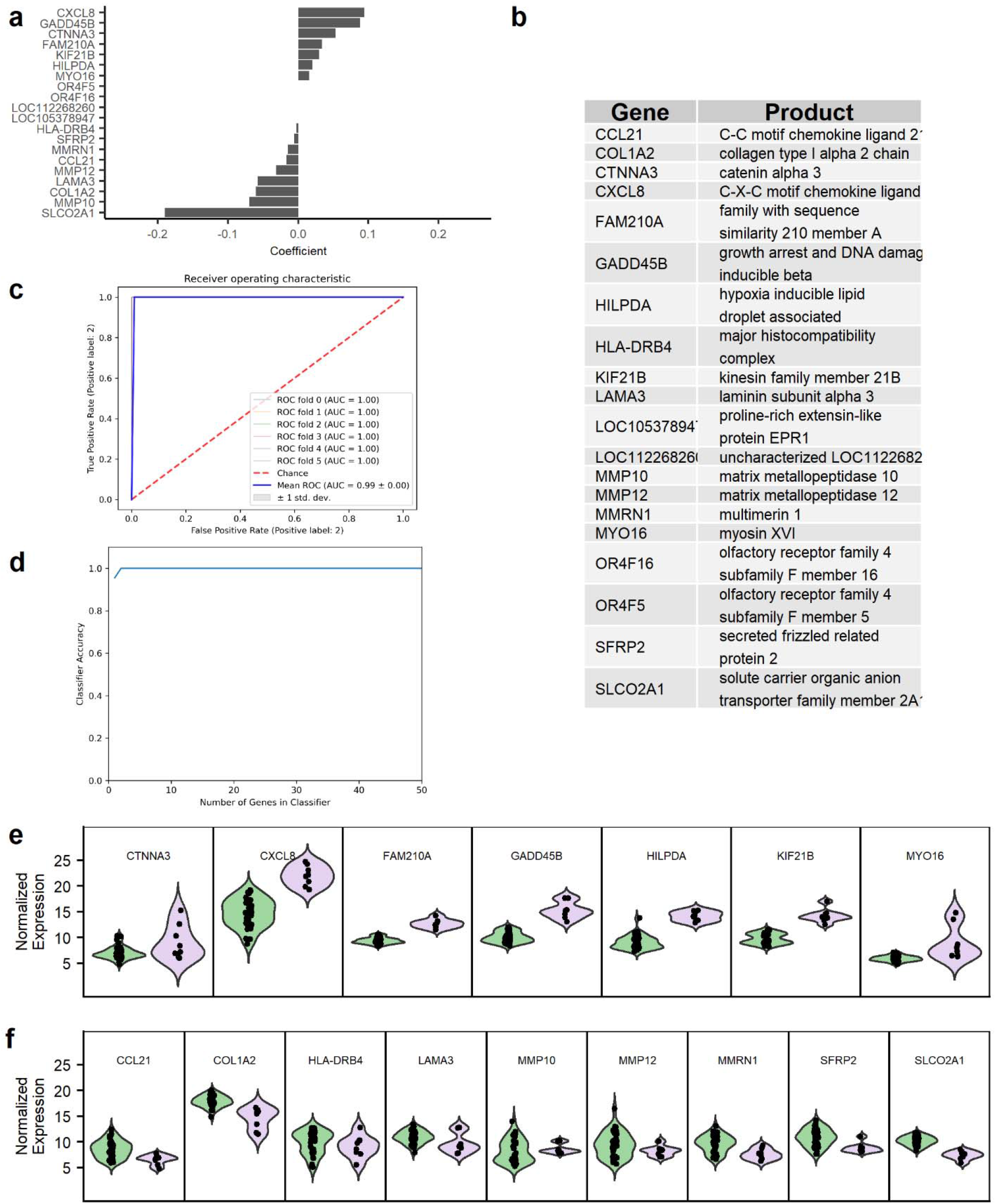
**(a)** Coefficient values for the top 20 genes extracted from the support vector classifier. **(b)** Gene symbols and products for identified features. **(c)** Receiver operating characteristic curve from cross-validation (6-fold, stratified) analysis. Given the 20 gene fingerprint, the classifier performed with 100% accuracy for classifying test samples in each fold. **(d)** Plot of classifier accuracy versus number of features included in the classifier and tested via stratified, 6-fold cross validation for each number of features.

We additionally performed the same analysis on the PEDIS/IDSA score to evaluate whether the model based on the clusters would perform to classify infection in samples compared to a model based on the clinical PEDIS/IDSA infection score classification system (Supplementary Figure 2,3). For the clinical PEDIS/IDSA, the model only achieved a maximum of 75-80% accuracy for predicting the PEDIS/IDSA score of the testing data with ∼30 genes (Supplementary Figure 4). This was also to be expected, as there did not appear to be a difference in gene expression for the features selected as good classifiers for PEDIS/IDSA score.

## Discussion

This study examined clinically infected DFUs to investigate whether clinical infection classification or ulcer duration reflect ulcers’ gene expression profiles, determined by RNA-sequencing. We observed that most of the variability in the data was not described by infection severity classification or ulcer duration. Rather, that the proportion of bacterial reads was a driving force in transcriptomic variation. Unsupervised clustering identified two groups of samples in the data, where one group demonstrated significantly increased bacterial activity. These groups were inconsistent with an ulcer’s infection classification and duration, despite the samples clustering in the same cluster. Samples with increased proportions of bacteria exhibited increased expression of genes associated with immune cells and inflammation, suggesting a direct response to the bacterial threat. We then identified a fingerprint of 20 genes, including molecules of the immune system such as *CXCL8*, a chemoattractant for neutrophils [26], which accurately identified samples exhibiting a transcriptome consistent with high proportions of bacterial RNA.

Developing a robust system for the stratification of ulcers is essential for treatment and study. Several studies have performed comparisons between ulcers, based on stratification by clinical classification, but often do not account for the influence of bacterial activity on the local microenvironment [21, 30-33]. For example, a previous study of infected ulcers identified increased expression of *GADD45B*, a DNA damage and stress response protein [27], (also identified in the present study) with IDSA/PEDIS scores of 4 [21].Our findings suggest rather that this gene and others are expressed only in a subset of samples with an IDSA/PEDIS score of 4, specifically those with high bacterial activity. Similarly, a previous study found no differences between microbiological data including presence of gram negative organisms or monomicrobial/polymicrobial infections among different grades of infection [33]. This is not to say that there is no difference in these parameters for severe infections, rather high variability among samples with the same grade may lead to decreased statistical power to detect these differences.

Our results suggest an acute inflammatory response to bacteria in C2 relative to lower-level inflammation observed in C1. This is supported by the inclusion of CXCL8 and other molecules induced by an inflammatory environment in C2, such as BCL2 Related Protein A1 (*BCL2A1*), oncostatin M (*OSM*), prostaglandin G/H synthase (*PTSG2*), and S-100 calcium binding protein A8/9 (*S100A8/9*). The increased expression of *CXCL8* and *ADAM8* also suggests active recruitment of neutrophils to the ulcer [34]. The presence of neutrophils in the samples is suggestd by the presence of hydroxycarboxylic acid receptor 2 (*HCAR2*) and oncostatin M (*OSM*), which are produced by or present on neutrophils [35, 36].. In addition, the increased expression of interleukin-6 (*IL6*) in C2 supports the presence of pro-inflammatory, type 1 macrophages. Contrarily, the presence of a reduced immune response and inflammatory environment was suggested in C1. For example, CXC13, a selective chemoattractant for B cells [37], showed increased expression C1 suggesting the recruitment of B cells. Previous research has suggested that the inhibition of immune response and recruitment of immune cells may lead to decreased wound healing, suggesting that C1 may represent a chronic-like state in comparison to C2, where we observed genes associated with acute inflammatory response and recruitment of neutrophils.

This study and the interpretation of the findings presents several limitations. First, distribution of bacteria is known to be heterogeneous within diabetic foot ulcers, but the heterogeneity of gene expression in an ulcer has not been studied. Thus, it is unclear whether the difference between clusters arises from global difference in wound gene expression or sampling regions with or without high levels of bacteria. We identified two samples in C2 with low bacterial activity, but yet clustered together with those exhibiting high bacterial activity, suggesting that the effect of bacteria may reach further than bacterias’ physical presence. This also further demonstrated the heterogeneity of bacterial distribution within wounds.

In summary, we identified that increased bacterial activity is associated with an inflamed and neutrophilic host transcriptome in lower extremity ulcers. The presence of bacteria also explained differences in host transcriptomes, despite equivalent clinical classifications. We then presented a small subset of genes, with consistent differential expression in samples with signs of bacterial infection, which proved to be accurate for identifying unknown samples with probably bacterial infections. Classification of wound status based on expression of important indicator genes may provide a more representative of the true wound environment than subjective clinical observations and may aid clinicians in determining the best course of treatment. Future work will be to improve the classification and identify the most relevant genes for describing the states of the wound. Our data reveals the potential future application of RNA-seq in profiling the metatranscriptome of DFUs based on their biological/cellular function. This may reduce any ambiguity of classifying wounds based on clinical observations and advance a more personalized medicine approach to care.

## Data Availability

All data produced in the present study are available upon reasonable request to the authors.

https://github.com/costerton-biofilm-center/WoundClassification2021

https://www.ncbi.nlm.nih.gov/bioproject/PRJNA726011

## Code and Data Availability

Raw sequence files for the RNA-seq data generated in this study are available at NCBI Sequence Read Archive (SRA) (Accession: PRJNA726011). The external data utilized in this analysis is available at NCBI SRA (Accession: PRJNA563930) [22]. Code to reproduce the analysis and figures in the manuscript is available at www.github.com/costerton-biofilm-center/WoundClassification2021.

## Ethics Statement

Study ethics approval was granted by the South West Sydney Local Health District Research and Ethics Committee (HREC/14/LPOOL/487).

## Contributions and Acknowledgements

MM, TB, and BF conceived and designed the project. MM collected and provided samples. BF Samples for this project were collected by MM. BF processed the samples. BF and JK performed the analysis. BF wrote the manuscript. BF, JK, CN, KK, MM, and TB read, revised, and edited the manuscript. Work for this project was supported by grants from The Lundbeck Foundation to TB and BF. TB is the guarantor of this work and, as such, had full access to all the data in the study and takes responsibility for the integrity of the data and the accuracy of the data analysis. We thank the GeoGenetics Sequencing Core at University of Copenhagen for assistance in generating the RNA-seq data and Computerome (www.computerome.dk) for providing the computing infrastructure to analyze the data. We thank Elio Rossi for useful discussions and contributions to the code used in the analysis. BF, JK, CN, KK, MM, and TB declare no conflicts of interest in relation to this work.

## Notes

### Competing Interest Statement

The authors have declared no competing interest.

### Funding Statement

Funding for this project was provided through grants awarded by The Lundbeck Foundation.

