## Supplementary Information for "Transcriptomic Fingerprint of Bacterial Infection in Lower Extremity Ulcers"

*Detailed description of Sample Collection for Liverpool Hospital (LHS) and Bispebjerg Hospital (BHS) data*

For the LHS samples, individuals presenting to a High-Risk Foot Service with an infected diabetic foot ulcer occurring below the malleolus were recruited over a twelve-month study period. A tissue punch biopsy was obtained from the edge of each DFU after debridement and cleansing of the wound with NaCl 0.9%. Tissue samples for RNA analysis were placed immediately into RNAlater® (Ambion, Inc) for 24 hours at 4°C and then frozen at -80°C until DNA extraction. Patients who received systemic antimicrobial therapy two weeks prior to enrolment were excluded. Patient demographics, laboratory and clinical data were collected through patient charts and the electronic medical records for correlation against RNA data. Clinical data and wound metrics of interest that were collected included; present or absent foot pulses, foot doppler waveforms, toe brachial indices (TBI) and completion of the modified neuropathic disability score. DFU location, duration of DFU prior to presentation, size (length x width in mm) and depth (mm). Laboratory data included; full blood count, inflammatory markers (White cell count [WCC], Erythrocyte sedimentation rate [ESR], C-reactive protein [CRP]), glycosylated haemoglobin (HbA1c) and estimated glomerular filtration rate (eGFR). All infected DFUs were diagnosed clinically, and their severity graded using the Infectious Disease Society of America Guidelines for DFI (Lipsky et al., 2012). Infection status (acute or chronic) were defined based on presenting clinical signs and symptoms in addition to duration. Study ethics approval was granted by the South West Sydney Local Health District Research and Ethics Committee (HREC/14/LPOOL/487). The study methodology conforms to STROME-ID and our molecular surveillance data are reported in keeping with this.

### **Supplementary Figures**

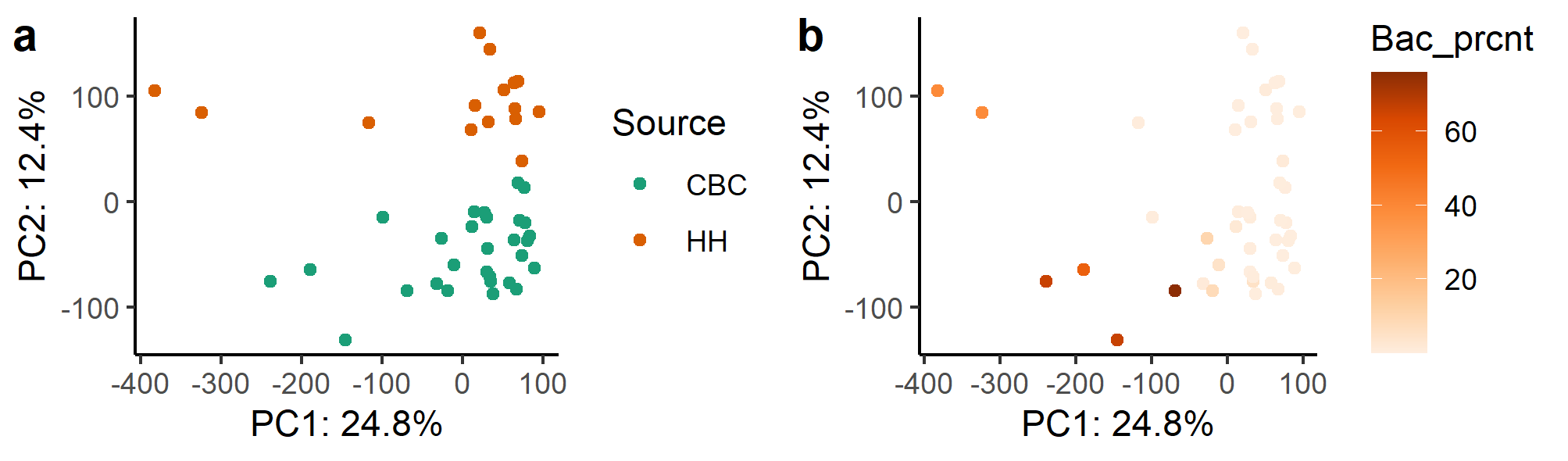

**Supplementary Figure 1.** PCA plot of vst-normalized count data prior to normalization for batch effects of (a) Source and (b) proportion of bacterial:human reads identified by RNA-seq.

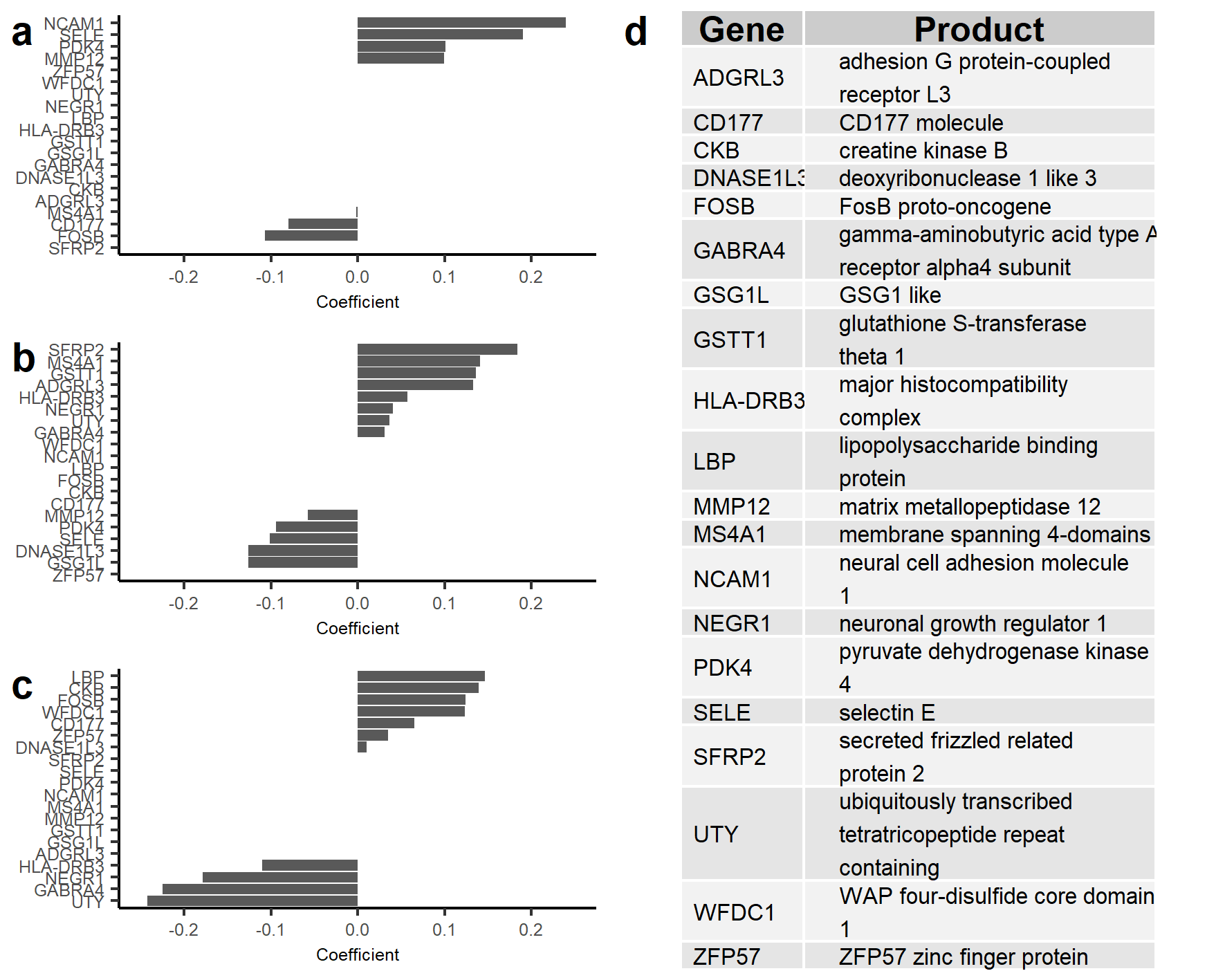

**Supplementary Figure 2. (a-c)** Coefficients for the twenty genes selected as a “fingerprint” for IDSA/PEDIS infection severity score for classification of **(a)** IDSA 2- mild, **(b)** IDSA 3- mild moderate, and **(c)** IDSA 4 - severe infections. **(d)** Gene symbols and product names for the twenty genes identified as the IDSA/PEDIS fingerprint.

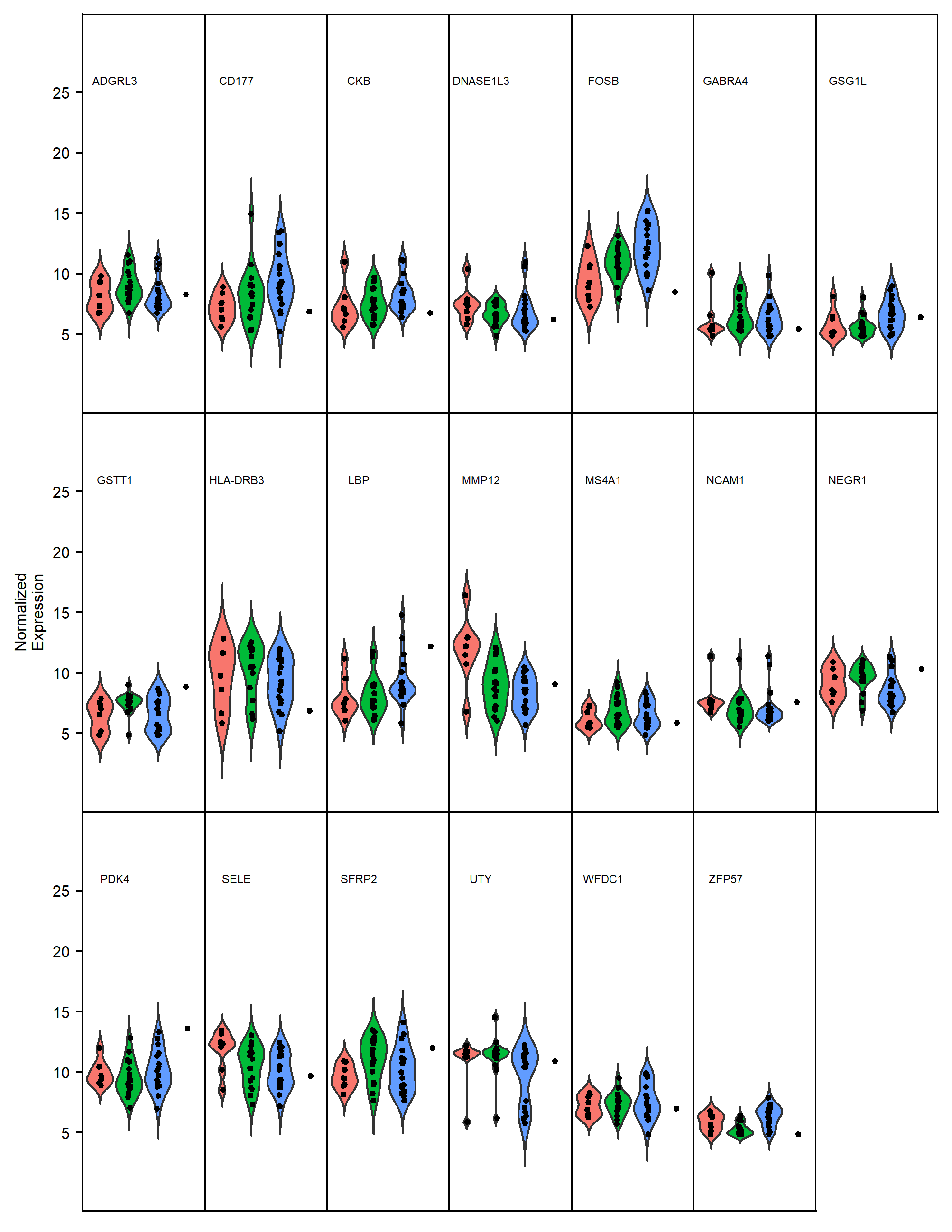

**Supplementary Figure 3.** VST-normalized gene expression values for the 20 genes selected to be effective classifiers of IDSA/PEDIS infection severity scores of 2(“mild”, red), 3(“moderate”, green), and 4 (s”severe”, blue). Samples missing IDSA/PEDIS values are displayed in the far-right group for each gene. Despite being classified as effective classifiers based on their coefficient weights in the SVC model, the expression of the majority of these genes appears similar between groups.

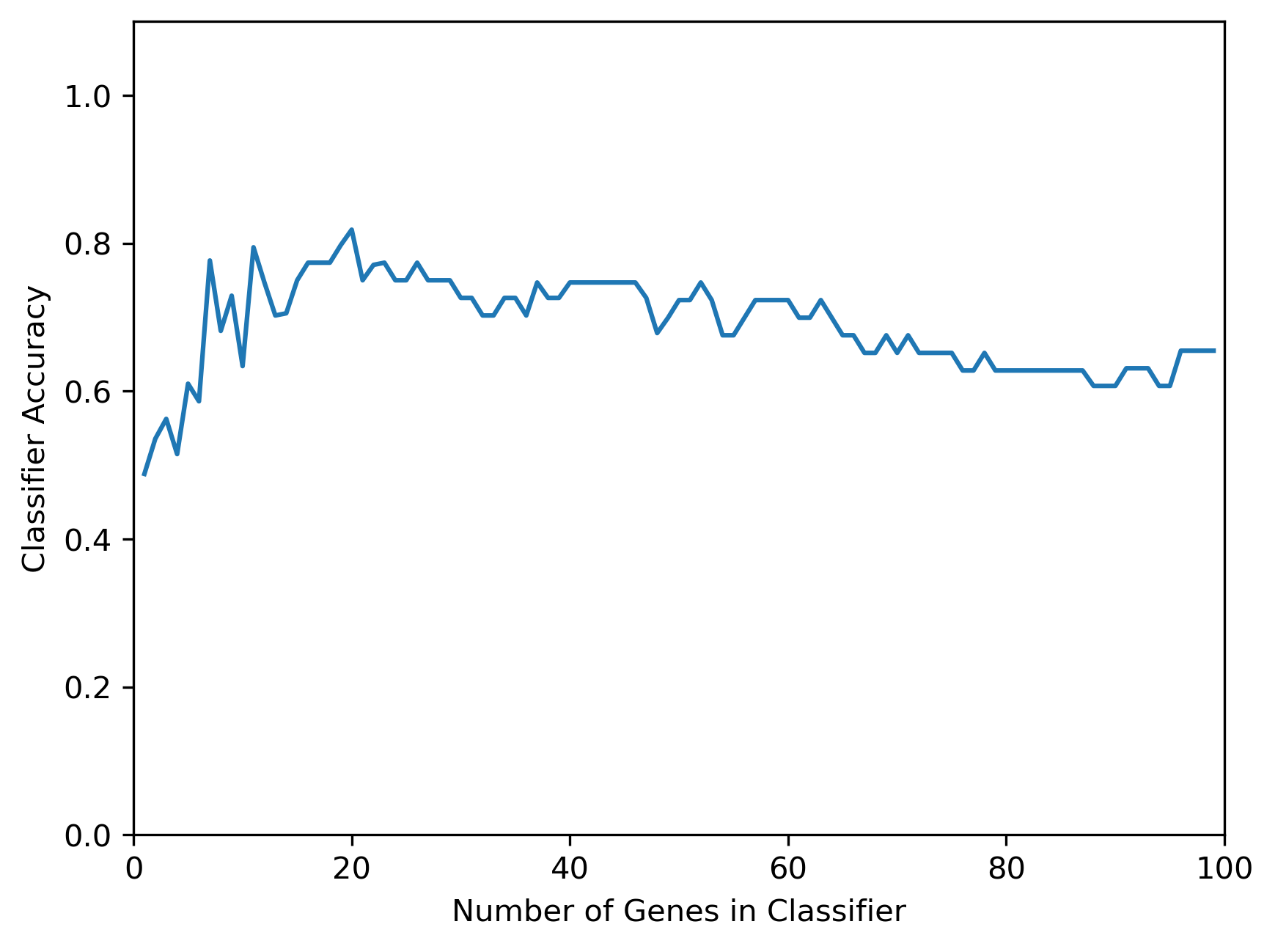

**Supplementary Figure 4.** Classifier accuracy for identifying IDSA/PEDIS scores of unknown samples with increasing number of gene-features included in the model. Accuracy was taken as the mean accuracy between folds based on a 6-fold stratified cross validation for each number genes.
